# Cohort profile: Description of the GIG-OSH longitudinal cohort on occupational safety and health of digital platforms workers in Europe

**DOI:** 10.64898/2026.03.05.26347679

**Authors:** Francesc Belvis, Edgar Vicente-Castellví, Sandra Verdaguer, Mariana Gutiérrez-Zamora, Joan Benach, Theo Bodin, Jessie Gevaert, Silvia Girardi, Jamelia Harris, Anna Ilsøe, Lauri Kokkinen, Trine P. Larsen, Sangwoo Lee, Filippa Lundh, Lluís Mangot-Sala, Nuria Matilla-Santander, Dorota Merecz-Kot, Hanna Nurmi, Chris Warhurst, Mireia Julià

## Abstract

**Purpose:** The GIG-OSH cohort was established to investigate the impact of digital platform work on occupational safety and health (OSH), working and employment conditions, and health in seven countries in Europe.

**Participants:** The cohort comprises 3,945 digital platform workers from seven European countries. The sample includes both web-based workers (e.g., micro-tasking, freelance design) and on-location workers (e.g., delivery, transport). Participants were recruited using non-probabilistic sampling strategies tailored to national contexts, including social media advertising, recruitment through micro-task platforms, and on-site field outreach. Multidimensional data have been collected through online surveys (implemented via REDCap) covering sociodemographic characteristics, working and employment conditions, psychosocial risks, algorithmic management, and physical and mental health indicators.

**Findings to date:** Participants had a mean age of 32.6 years at baseline (SD 10.4), and the majority are male (58.8%), with a higher concentration of migrants in on-location tasks (62.2%) compared to web-based tasks (48.8%). Regarding educational attainment, 55.4% of the total cohort holds a tertiary degree, reaching 64.4% among web-based workers. Platform work intensity varies significantly: on-location workers averaged 85.4 hours of work in the last month, while web-based workers averaged 47.0 hours. Mean income from platform work as a percentage of the national median was 20.6% (SD 22.2). The mean WHO-5 Well-Being Index score was 58.7 (SD 20.3), which is notably lower than the European general population average (69.4), indicating poorer mental health outcomes among cohort members.

**Future plans:** The GIG-OSH cohort represents the first large-scale, longitudinal study examining occupational safety and health among digital platform workers across multiple European countries. Future waves will prioritize developing precise tools to measure hourly earnings and unpaid waiting time. Future research should aim to include underrepresented subgroups, such as medical and domestic care workers, and explore potential linkage with administrative records to evaluate long-term health trajectories and the impact of new EU labour regulations.

**Strengths and limitations of this study:**

- This is the first large-scale longitudinal cohort to examine occupational safety and health among platform workers across multiple European countries, addressing an important evidence gap.
- The inclusion of both web-based and on-location workers enables comparative analyses across diverse task types, employment conditions, and national contexts.
- Recruitment strategies tailored to national contexts enhanced feasibility but limited the representativeness of samples and precluded national-level weighting or benchmarking.
- High attrition between waves and the absence of harmonized classifications (e.g., education levels) across countries may constrain the generalizability and longitudinal consistency of findings.
- Despite relying on self-reported data, the study used stakeholder-informed instruments and captured a wide range of occupational hazards—such as psychosocial and algorithmic risks—not typically addressed in conventional labour surveys.

## INTRODUCTION

In recent years, digital platform work (DPW) has strongly emerged onto the European labour market, introducing new forms of employment characterised by algorithmic management and task-based remuneration via apps (1). This trend has accelerated in the wake of the COVID-19 pandemic, which catalyzed the expansion of platform-based services, including food delivery, transportation, and digital microtasks (2). Recent EU pilot data further highlight the consolidation of this phenomenon: in 2022, 3.0% of people aged 15–64 in 17 European countries reported having engaged in digital platform work within the last 12 months, but different national accounts point to wide cross-country variations – UK 1,4% to Spain 4,3%. The majority of those workers were concentrated in younger cohorts and among tertiary-educated workers. Digital platform workers can be roughly divided into three main parts: young people aged 15–29 (30.6%), females aged 30–64 (32.8%), and males aged 30–64 (36.6%) (3). This form of employment spans diverse sectors, and it can be broadly categorized into: a) web-based platform work, which distributes tasks to a geographically dispersed workforce through open calls, and b) on-location platform work, which refers to location-specific applications that assign tasks to workers within defined geographic areas (5,6). It should also account for the wide variability in platform tasks, ranging from short, low-intensity click work to longer-term but fixed projects, and involving markedly different levels of skill requirements.

Despite the promise of increased flexibility (6), platform work often entails precarious conditions, such as low salaries, temporality, lack of training, no guaranteed minimum working hours, unpaid work, no health or retirement benefits and limited social protection (7). These conditions are particularly concerning in light of the growing body of evidence linking precarious work to adverse mental health outcomes (8,9). A recent Belgian study by (10) revealed that they experienced a lack of workplace rights, economic unsustainability, undesirable working times, and weak collective organisation. These factors were significantly and positively associated with poor well-being (10). In a study conducted in Italy on riders, it was found that this group is exposed to a high risk of experiencing road accidents or violence by third parties (11). Although research on platform work is growing, few studies address occupational safety and health (OSH) and many are country based qualitative studies, not comparative nor quantitative. A recent scoping review highlights these gaps and the lack of longitudinal data on OSH impacts of digital platform work (12). Despite the previous studies, longitudinal data examining the occupational safety and health (OSH) impacts of digital platform work remain scarce (12).

This gap is particularly concerning given the previously mentioned heterogeneity within the gig economy, where workers perform a wide range of tasks under diverse conditions that may influence their physical and mental health differently over time. For example, on-location settings, such as food delivery platforms, often involve a significant presence of migrant workers (11), while web-based platform work tends to attract women, young individuals, and those who seek flexibility, income supplementation, or access to wider job markets (6). These differing contexts may cause variability in health outcomes among platform workers.

Due to this, understanding its implications for social determinants of health has become crucial. To address these gaps, the GIG-OSH cohort, which represents one of the first large-scale comparative efforts to collect different point-in-time data, was established to investigate the health-, safety-, working-, and employment conditions of digital platform workers across seven European countries (Belgium, Denmark, Finland, Poland, Spain, Sweden and the UK), each representing distinct welfare settlements with different industrial relations systems for regulating wage and working conditions (13). This cohort is part of the GIG-OSH project that adopted a holistic approach to the study of platform work by integrating quantitative and qualitative methods. Beyond combining methodological perspectives, the GIG-OSH project explicitly incorporates the viewpoints of platform workers as well as those of key stakeholders and institutional actors. The broad research questions of the GIG-OSH cohort are: (1) How do the working conditions and employment arrangements of digital platform workers across different European countries influence their occupational safety, health, and well-being over time? (2) How do working, employment conditions, and occupational risks differ between web-based and on-location platform workers in different European countries? Building on these overarching research questions, the objective of this paper is to present the GIG-OSH cohort profile, which addresses the current lack of cross-national, longitudinal data on occupational safety and health (OSH) among platform workers. Specifically, it aims to describe demographic, task types, employment conditions and exposures, physical and mental health, and mental well-being outcomes. It also provides a multidimensional description of the relationship between the type of tasks performed under digital platform arrangements and the main employment conditions under which these tasks are developed. The paper is structured as follows: Section 2 describes the cohort design, recruitment, data collection, variable definitions, and statistical analysis; Section 3 reports findings to date; Section 4 provides a multidimensional description of task types and employment conditions; Section 5 discusses strengths and limitations; and Section 6 the conclusions.

## COHORT DESCRIPTION

### Study and questionnaire design

The GIG-OSH study employs a prospective longitudinal cohort design to investigate occupational safety and health (OSH) among digital platform workers across Europe. The study comprises a baseline survey and one follow-up wave at six months, with the exception of Spain, where a second follow-up wave at one year was also conducted.

The baseline survey was conducted between October 2023 and April 2024 across seven European countries through a collaboration involving nine academic institutions: Belgium (Vrije Universiteit Brussel and Université libre de Bruxelles), Denmark (University of Copenhagen), Finland (Tampere University), Poland (University of Lodz), Spain (Hospital del Mar Research Institute and Pompeu Fabra University), Sweden (Karolinska Institutet), and the United Kingdom (University of Warwick).

Data collection was conducted using online questionnaires tailored to the specific working conditions and occupational risks experienced by platform workers. The survey instrument was developed based on a comprehensive review of existing literature and the research team’s prior experience with non-standard and platform-based labour studies. To ensure relevance and validity, a co-creation process was employed that incorporated input from collaborating researchers, partner organisations, and stakeholders involved in platform work advocacy, policy, and research. The questionnaire was translated into the relevant languages of each participating country using a standardized translation process and was subsequently pilot tested before fieldwork. Data collection was conducted through the REDCap application. The survey included multiple sections covering demographic details, work conditions, income levels, experiences with platform work, and occupational safety and health. To improve response quality, an attention check question was incorporated throughout the survey. Additionally, time-tracking measures were implemented to identify and filter out responses that were completed too quickly to be reliable.

The second wave of data collection was launched six months after the baseline and included all participating countries. A third wave, conducted exclusively in Spain 12 months post-baseline, was made possible due to the particular interest expressed by the Spanish research team, as well as the availability of both human and financial resources to support extended follow-up in this national context. This third wave was not included in the multidimensional analysis presented in the subsequent sections of this paper.

### Recruitment strategies

For the purposes of the GIG-OSH study, digital platform workers were considered individuals who earn income by providing labour or services through digital platforms. In line with EU-OSHA (14), these platforms act as intermediaries, typically mediated by algorithms and online interfaces. Following the ILO’s classification (15), platform work in the study includes both on-location (e.g., ride-hailing, food delivery, household services) and web-based work (e.g., online freelancing, microtasks). In contrast, web-based platform work involves tasks that are performed entirely online and are not tied to a geographic location, such as data entry, translation, design, online teaching, large IT project, or other freelance digital services. To be eligible for inclusion in the GIG-OSH cohort, participants had to report currently earning money, either regularly or occasionally, by providing goods or services through a digital platform, website, or application, and reside in one of the seven participating countries. This definition was intended to reflect the heterogeneity of platform work across sectors, work arrangements, and national contexts. It focuses specifically on labour that is mediated and organized through digital infrastructures, regardless of the worker’s formal employment status or legal classification.

The initial target sample was 500 digital platform workers per country, evenly distributed across the two modalities. To ensure broad and diverse participation across both on-location and web-based segments of the platform workforce, each country implemented a purposive sampling approach (16). The methods were tailored to national contexts while maintaining common objectives of reaching hard-to-access groups and ensuring data quality. These approaches combined different strategies used by each country, summarized in Table 1:

**Table 1.** The enrolment approaches and follow-up contact procedures used by each country. An “x” indicates that a given strategy was implemented in that country. Strategies shaded in light grey indicate those that were less effective, while those shaded in dark grey represent the approaches that had the greatest impact on participant enrolment.

| COUNTRIES | Belgium | Denmark | Finland | Poland | Spain | Sweden | The UK |
| --- | --- | --- | --- | --- | --- | --- | --- |
| ENROLMENT APPROACHES |  |  |  |  |  |  |  |
| 1. Targeted Digital Advertising (Social Media Ads) | x | x | x |  | x | x | x |
| 2. Organic Outreach via Social Media Groups and Networks | x | x | x | x | x | x | x |
| 3. Microtask Platforms (Upwork and Prolific) | x | x | x |  | x | x | x |
| 4. Fieldwork and In-Person Recruitment | x | x | x |  | x | x | x |
| 5. Institutional Collaboration (Unions, Universities, Agencies, Platforms) | x | x | x |  | x | x | x |
| 6. Snowball Sampling and Community-Based Strategies |  |  |  | x | x | x |  |
| 7. External Research Agency Support |  |  |  | x |  |  |  |
| 8. Recontacting Previous Study Participants | x |  |  |  |  | x |  |
| FOLLOW-UP CONTACT PROCEDURES |  |  |  |  |  |  |  |
| Three standardized email invitations sent via REDCap at three-day intervals | x | x | x | x | x | x | x |
| Two additional personalized reminder emails sent directly by the researcher |  |  | x |  |  |  |  |
| Direct WhatsApp messages sent to participants who had provided mobile numbers |  |  |  |  | x |  |  |
| Follow-up survey posted as a microtask on Prolific |  |  |  |  | x | x |  |

#### 1. Targeted Digital Advertising (Social Media Ads)

This method involves the use of paid advertisements on social media platforms such as Facebook, Instagram, X (formerly Twitter), and LinkedIn to reach potential participants, who are likely to be platform workers. Ads were carefully tailored in content and visuals to appeal to specific worker groups (e.g., couriers, drivers, freelancers) and were geographically or demographically targeted to increase relevance and engagement. In some cases, ad performance was monitored and adjusted throughout the campaign to improve reach and cost-efficiency.

#### 2. Organic Outreach via Social Media Groups and Networks

This approach leveraged existing online communities where platform workers interact, such as Facebook groups, Telegram channels, and LinkedIn networks. Instead of using paid promotion, researchers posted survey links organically and engaged directly with users. In some countries, this included identifying and contacting influencers or individuals who post frequently about gig work (e.g., on TikTok, Instagram), inviting them to participate or share the survey.

#### 3. Microtask Platforms (Upwork and Prolific)

Enrolment was also conducted by posting the survey as a paid assignment on platforms like Upwork, which caters to freelancers, and Prolific, which connects researchers with potential respondents. This method was especially useful for reaching web-based platform workers, such as freelancers and micro-taskers, who may not be accessible via traditional outreach. It also allowed financial compensation, which improved completion rates and attracted a more diverse pool of workers.

#### 4. Fieldwork and In-Person Enrolment

This strategy involved physical presence in urban areas, transport hubs, or other locations where platform workers (especially couriers and drivers) tend to gather. Researchers distributed flyers, engaged in direct conversations, and sometimes attended relevant events or forums. This face-to-face contact allowed enrolment of hard-to-reach groups, particularly those not active on social media or online platforms. Fieldwork also helped validate participants’ platform work and improve inclusivity.

#### 5. Institutional Collaboration (Unions, Universities, Agencies, Platforms)

Researchers collaborated with trade unions, student organisations, public agencies, and platform companies to distribute the survey through established communication channels such as email lists, newsletters, internal dashboards, and social media pages. This method leverages the credibility and reach of established institutions, helping to access trusted communities and boosting the legitimacy of the study. In some cases, tailored materials were created to facilitate internal promotion.

#### 6. Snowball Sampling and Community-Based Strategies

This method relied on peer-to-peer referrals; participants were encouraged to invite others in their network who also work on platforms. In some cases, this was combined with word-of-mouth strategies or engagement through local worker networks or informal collectives.

#### 7. External Research Agency Support

In cases where enrolment via typical academic channels proved insufficient, external market research agencies were brought in to assist with outreach. These agencies often have expertise in targeting specific demographics, access to respondent databases, and experience with online and offline campaign management. Their support enhanced sample diversity and response rates, especially in challenging or low-trust contexts.

#### 8. Recontacting Previous Study Participants

This strategy involved inviting individuals who had already participated in an earlier, related study (16) to take part in the current study. Re-engaging this qualified sample improved efficiency and enabled potential longitudinal insights, while fully complying with GDPR regulations and not compromising the study’s ethics approval.

### Follow-up Contact Procedures

To support the second wave phase of data collection, all participating countries implemented a standardized recontact protocol using the REDCap platform. Participants who had previously consented to be recontacted and had provided valid contact information received three email invitations, spaced at three-day intervals.

In addition to the standardized REDCap email outreach, some country teams implemented supplementary contact strategies to enhance response rates and participant engagement (Table 1). In Finland, for example, two personalized reminder emails were sent directly by the researcher to each participant who had provided valid contact information. In Spain, additional strategies included direct WhatsApp messages sent to participants who had provided mobile numbers, as well as, also in Sweden, recontact through online platforms such as Prolific, where the follow-up survey was posted as a microtask.

### Data collection and participants

The data collection process was designed to ensure the accuracy and reliability of responses while maintaining ethical research standards. Surveys were administered online through a secure survey platform (REDCap), with participants recruited using the strategies outlined above. For in-person enrolment, field researchers assisted participants in survey completion where necessary, ensuring clarity in responses. All data was anonymized and securely stored.

The collected data was systematically cleaned and analyzed, with missing or inconsistent responses reviewed before finalizing the dataset. These measures ensured high-quality data, allowing for meaningful analysis of platform work conditions across the different countries included in the study.

Figure 1 describes the flow of participants in the process of assessing eligibility, recruitment, and follow-up. At baseline, 8067 individuals accessed the survey through the REDCap system. After 2,089 exited before consent and 29 declined, 5,952 participants provided informed consent. Following self-eligibility screening, 5 individuals were excluded for residing outside the seven countries included in the study. Additionally, 620 participants were excluded for not being platform workers (597 did not earn income through platforms, 14 did not respond, and 9 were uncertain and did not select any platform). As a result, 5,327 individuals passed the self-eligibility screening and completed the online survey. During data cleaning, 1,382 additional exclusions were made: 130 duplicate entries and 1,252 individuals who did not meet the investigator-defined platform worker criteria (e.g. apps not related to DPW). This resulted in a final baseline cohort of 3,945 participants.

**Figure 1.**
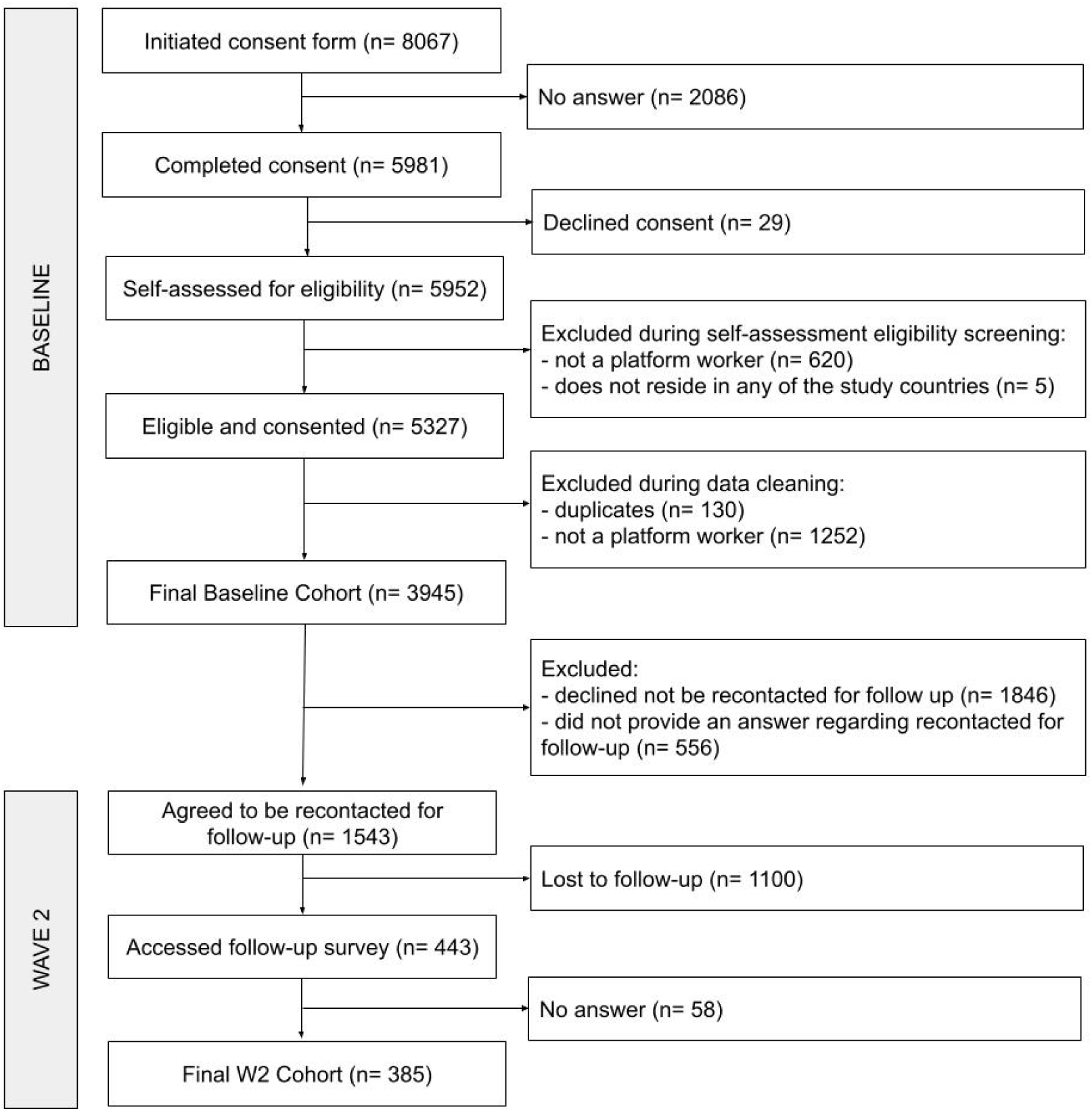
Flow diagram of the GIG-OSH participants for baseline and wave 2.

For Wave 2 (see also Figure 1), all 3,945 baseline participants were considered. However, 1,846 had declined to recontact at baseline, and 556 did not respond to the recontact consent question, leaving 1,543 participants who agreed to follow-up. Among them, 1,100 were lost to follow-up, resulting in 443 individuals who accessed the Wave 2 survey. Of these, 58 exited without proceeding, yielding a final Wave 2 cohort of 385 participants.

### Variables definition/values

The questionnaire collected detailed information on the occupational hazards and working conditions experienced by digital platform workers. These included technology-related risks (e.g., technostress, cyberbullying), criminal risks (e.g., exposure to sexual harassment), and traffic-related risks (e.g., road safety concerns, hazardous weather conditions). Psychosocial risks were also examined, such as discriminatory behaviour from clients, fear of receiving negative customer ratings, and the availability of dispute resolution mechanisms. Additionally, the study assessed risks associated with algorithmic management, including stress related to work allocation, performance monitoring, and evaluation processes.

The questionnaire also collected information related to working hours (e.g., unpredictability, poor work-life balance), tenure as a platform worker, and employment arrangements (e.g., self-employment, direct employment, hybrid or multiple job holding) and earnings. Furthermore, the study explored the use of personal protective equipment (PPE), such as helmet usage, as well as access to training programs, including instruction on PPE use and compliance with traffic regulations. Self-reported health status was assessed, with a particular focus on mental health and general health perception as well as mental well-being that was assessed by WHO-5 (17). Finally, sociodemographic characteristics such as gender, age, educational attainment, country of birth, and immigration status were recorded to provide a comprehensive understanding of participants’ characteristics.

To ensure comparability across different aspects of working conditions, we constructed composite indices using standardised measures. The indices for working time quality, work intensity, physical environment, autonomy, and social environment were derived from items of the European Working Conditions Survey (EWCS), in line with the official Eurofound methodology (18). Each index ranges from 0 to 100, with higher values indicating more favourable conditions, except for the work intensity index, where higher values denote greater intensity. In addition, we developed a contract type index (0–100), capturing the degree of employment formality, and an earnings index (0–100), reflecting normalised hourly wages relative to the country-specific median. See table SA1 in supplemental material A to know the variables included in each index.

For the multivariate analysis of the relationship between DPW tasks and employment conditions, we employed two sets of variables in the GIG-OSH database: (a) ‘main task’ (a list of fifteen specific tasks, see table SB1 in supplemental material B) and (b) employment conditions, including ‘Type work arrangement’ (Self-employed, Permanent, Fixed-Term, Student Contract, No Contract, Other/DK Contract), ‘Doing another job? (Y/N)’, ‘Income from platform work as a percentage of median country income’, ‘Monthly time commitment (in hours)’, and ‘Share of personal income from DPW (%)’ (the last three variables recoded into quintiles). The worker’s country of residence and a binary recoding of the task type (web-based/on-location) were also included as supplementary variables.

### Statistical analysis

The statistical analysis plan was designed to describe the collected data and identify key associations within the cohort. Descriptive analyses were conducted to characterize our population cohort with a focus on demographic variables (e.g., age, gender, educational level), and occupational variables (e.g., type of platform, working hours, earnings). For continuous variables, measures of central tendency (mean) and dispersion (standard deviation) were calculated. For categorical variables, frequencies and percentages were reported. To examine associations among variables and uncover latent structures, we also employed nonlinear canonical correlation analysis, a multivariate technique suitable for multiple sets of variables measured at differing levels (19). After excluding 820 cases with missing data, the effective sample comprised n = 3,125. The centroids of the variable categories were projected onto a biplot based on the first two dimensions extracted by the analysis, allowing visualization of the main patterns of association. The worker’s country of residence and the web-based versus on-location nature of the task were included as non-active supplementary variables.

All the analyses were performed within the R statistical environment (v4.4.1; (20)). The *homals* package v.1.0.10 (21) was used for non-linear canonical correlation analysis, and the ggoplot2 package v. 3.5.2 for Figure 2.

**Figure 2.**
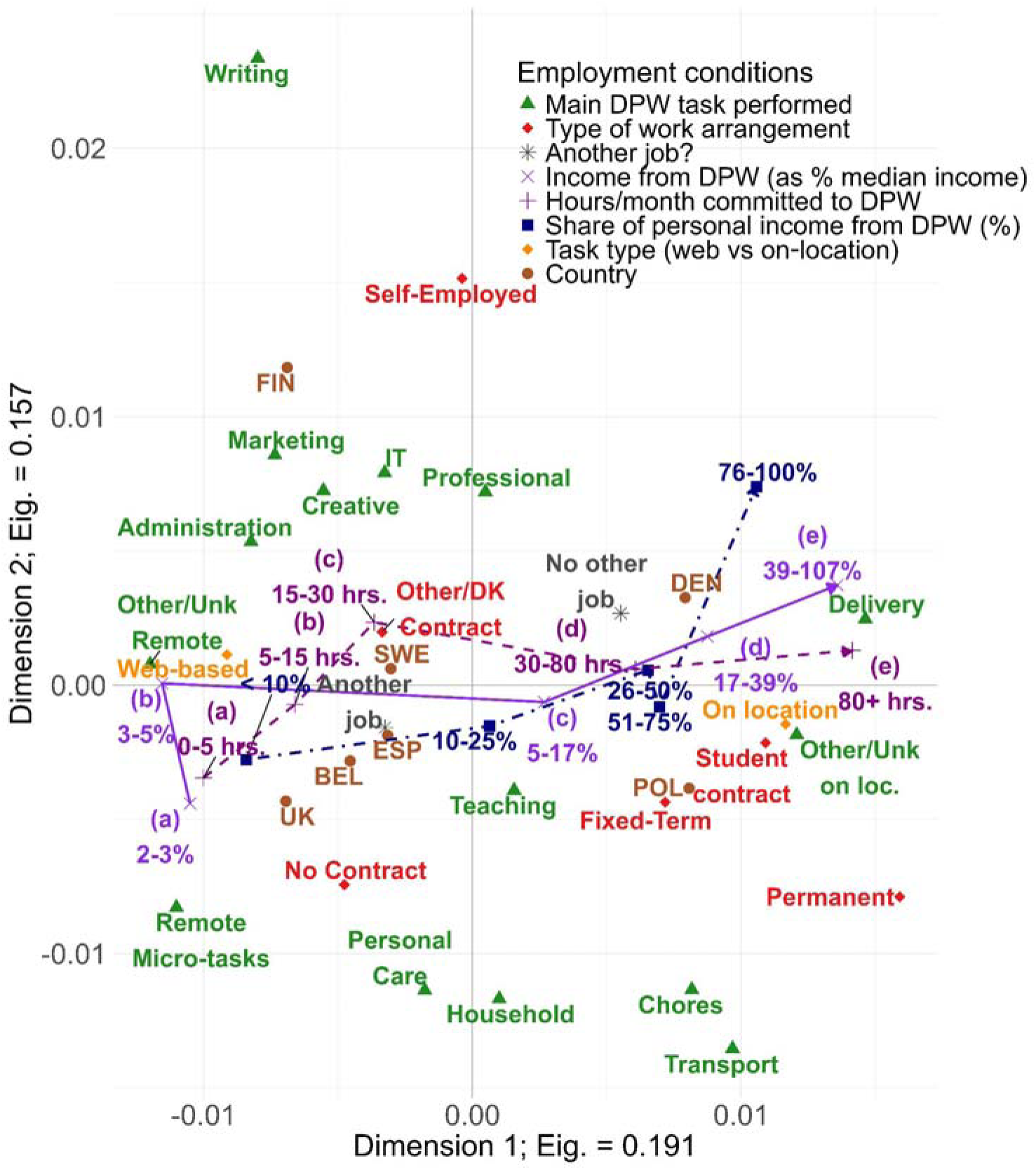
Joint category quantization plot (centroids) for the first two dimensions of the 3-D HOMALS solution.

## FINDINGS TO DATE

### Sociodemographic and labour characteristics

Baseline sociodemographic and labour characteristics of the 3,945 participants in the GIG-OSH cohort are presented in Table 2. Participants were stratified by type of platform work-on-location versus web-based- to examine subgroup-specific patterns. Gender distribution was balanced among web-based workers (48.84% male), whereas men were overrepresented in on-location work (69.16%). Differences were also observed in migration background, with a higher proportion of immigrants among on-location workers (41.17%) compared to web-based (27.04%). Educational attainment was notably higher among web-based workers, 64.42% of whom had completed tertiary education, compared with 45.94% in the on-location group.

**Table 2.**
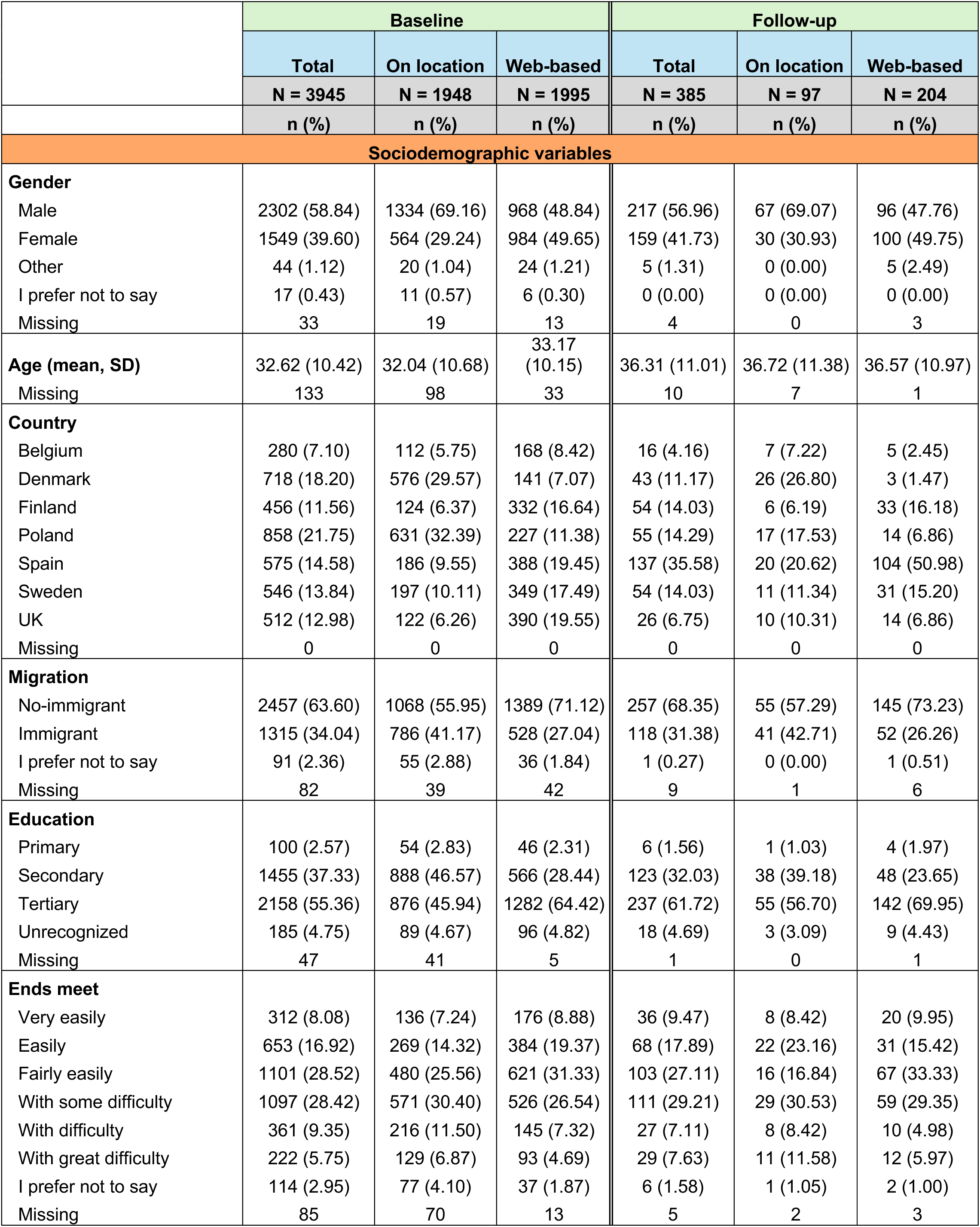
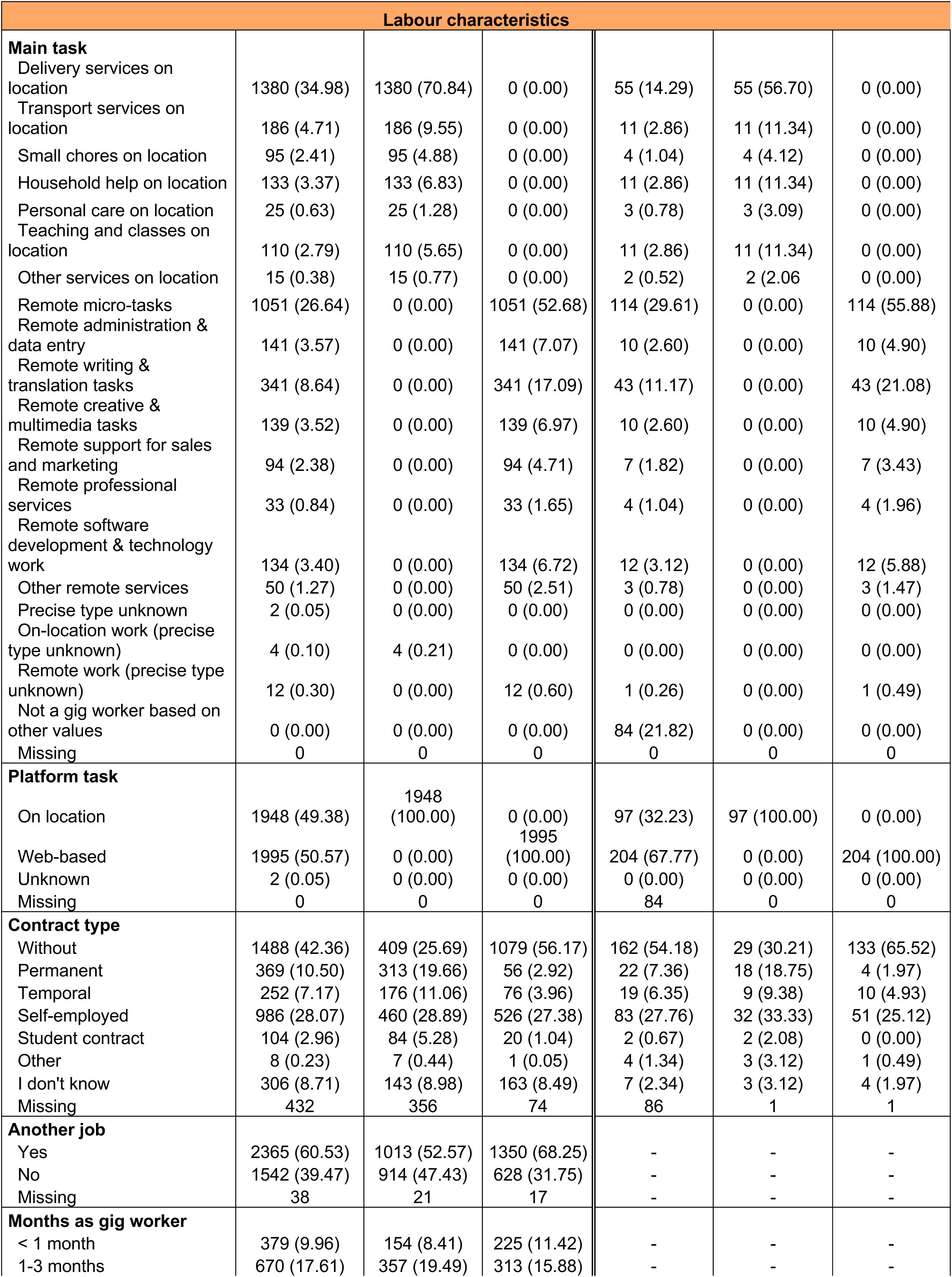
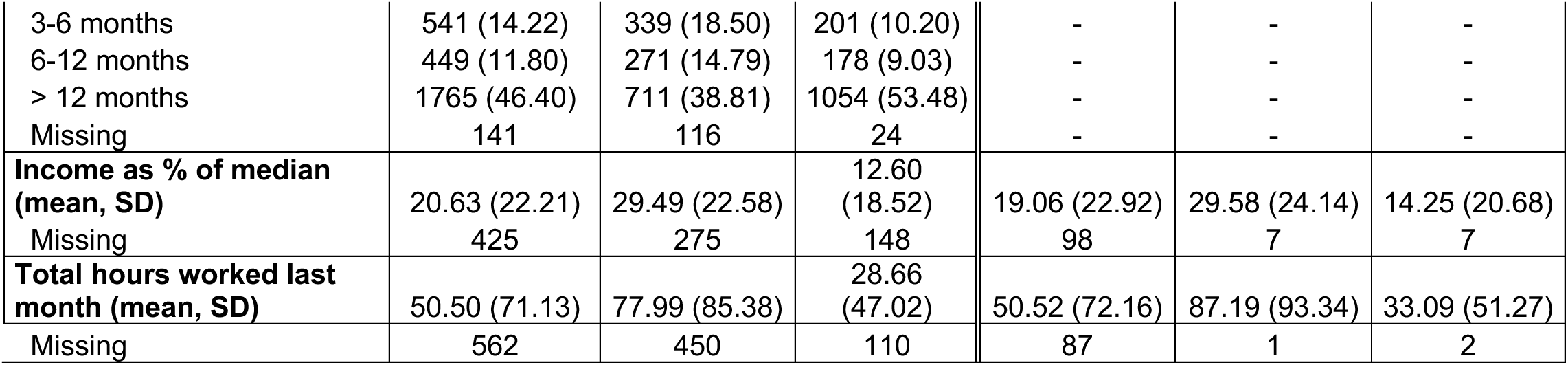
Baseline (n=3,945) and follow-up (n=385) sociodemographic and labor characteristics of the participants in the GIG-OSH cohort.

Substantial differences were also observed in professional characteristics in the baseline (Table 2). Web-based workers were primarily engaged in micro tasks (52.68%), while the majority of on-location workers were involved in delivery services (70.84%). Employment arrangements varied considerably: 56.17% of web-based workers reported having no formal employment contract, compared with 25.69% among on-location workers. Although holding another job was common across both groups, prevalence was higher among web-based workers (68.25% vs 52.57%). A greater proportion of web-based workers reported more than 12 months of experience in platform work (53.48%) compared to on-location workers (38.81%). In terms of income, earnings were generally low in both groups; on average, web-based workers reported earning only 12.60% of the national median income, while on-location workers earned 29.49%. Finally, web-based workers reported an average of 28.66 working hours per month, whereas on-location workers reported a mean of 77.99 hours.

Sociodemographics and labour characteristics, available for the subsample of 385 participants in follow-up, are also presented in Table 2. Stratified results by country for both baseline and follow-up assessments are provided in Supplemental Material Table SA2.

### Occupational safety and health, self-perceived health and work environment characteristics

Baseline occupational safety and health (OSH) conditions, self-perceived health, and work environment characteristics are presented in Table 3, stratified by type of platform work (on-location vs. web-based). In terms of OSH information, a slightly higher percentage of on-location workers felt well-informed (45.27%) compared to web-based workers (40.96%). However, web-based workers were more likely to report not feeling informed at all (12.84%) compared to on-location workers (8.91%). Clear contrasts were observed in the availability of protective equipment. Among on-location workers, 19.83% reported receiving some kind of equipment paid by the platform, while among web-based workers this figure was below 3%. Notably, 74.12% of web-based workers indicated not using any protective equipment, versus 29.34% among on-location workers.

**Table 3.**
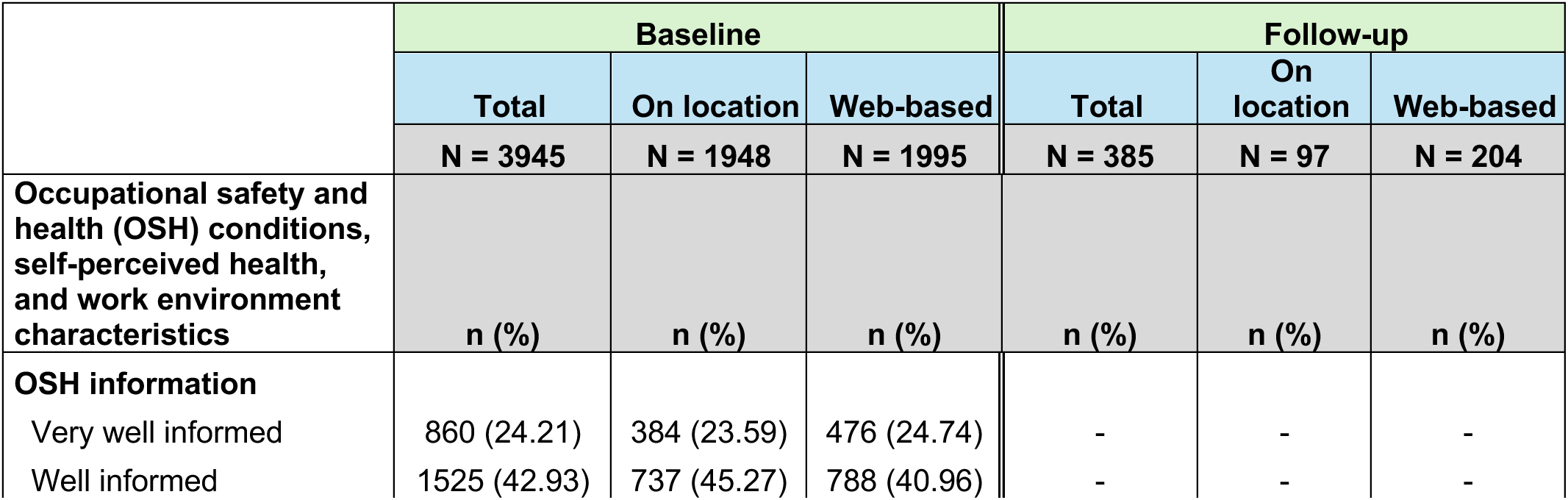
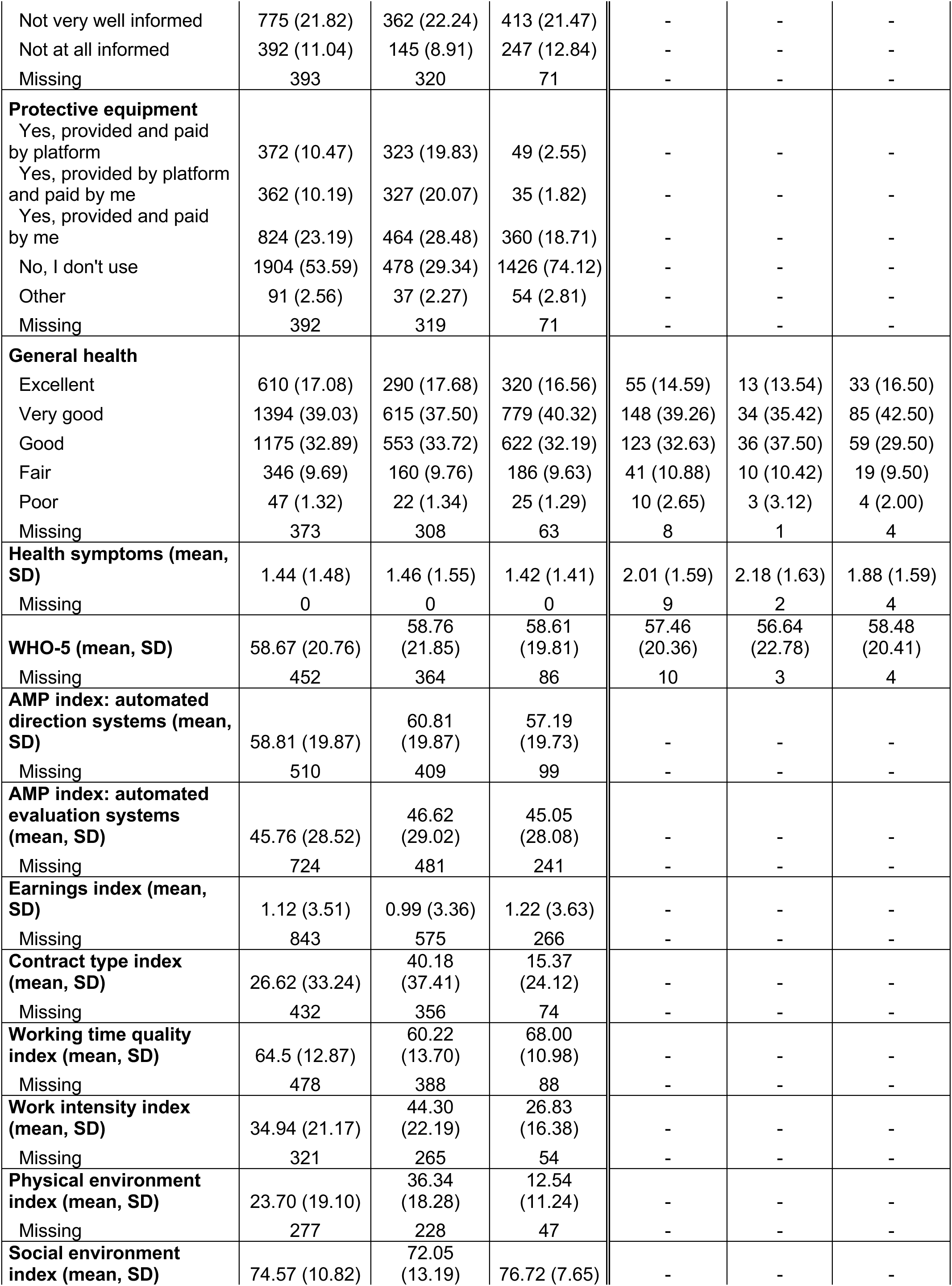
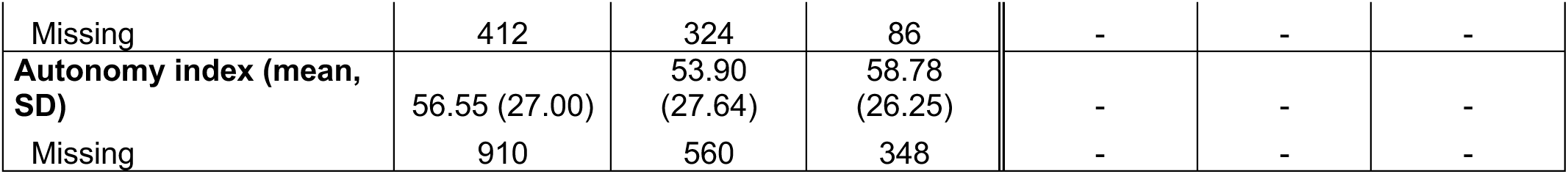
Baseline (n=3,945) and follow-up (n=385) occupational safety and health (OSH) conditions, self-perceived health, and work environment characteristics of the participants in the GIG-OSH cohort.

Self-perceived general health at baseline was rated as very good or excellent by a majority of participants in both groups. The average number of self-reported health symptoms was 1.44 (SD=1.48) on a 0-9 scale. The psychological well-being, assessed via the WHO-5 index, showed similar scores across groups (mean: 58.67, SD=20.76). This score is lower than the EU-wide mean of 69.4 according to the 2024 EWCS (18), indicating poorer psychological well-being in our sample.

Working conditions varied considerably by type of platform work. The earning index was low in both groups, with slightly higher values among web-based workers (1.22 vs. 0.99). The contract type index was higher among on-location workers (40.18 vs. 15.37), suggesting more formal employment arrangements, as shown in Table 2. On-location workers reported lower working time quality (60.22 vs. 68.00), higher work intensity (44.30 vs. 26.83), and a more demanding physical environment (36.34 v 12.54). In contrast, web-based workers reported better-rated social environments (76.72 vs. 72.05) and greater autonomy (58.78 vs. 53.90).

Follow-up data generally confirmed patterns observed at the baseline, though some indicators suggested a deterioration in perceived health, but not in psychological well-being (Table 3). Stratified results by country for both baseline and follow-up assessments are provided in Supplemental Material A, Table SA3. Nevertheless, these results should be interpreted with caution, given the substantial attrition rate observed at follow-up.

### Multidimensional Description

This section presents the results of the nonlinear canonical correlation analysis examining the relationship between the types of tasks performed in digital platform work and the principal employment conditions under which they are carried out. This approach provides a comprehensive perspective on the key associations within the data. Here, we provide a concise summary of the main findings, while full methodological details are presented in Supplemental Material B.

The extremely low loss value (0.0000131) of the procedure indicated minimal information loss during the optimal scaling phase. A three-dimensional solution was extracted using homogeneity analysis (HOMALS) based on optimally scaled variables, accounting for comparable proportions of variance across dimensions (D1 = 0.191, D2 = 0.157, D3 = 0.155). Figure 2 presents the first two dimensions derived from these computations. Dimension 1, represented on the X-axis, primarily distinguishes between web-based and on-location tasks, along with their associated differences in key employment conditions. On the left side of this axis are clustered remote forms of work, including micro-tasks, writing, marketing, and creative activities. These tasks, particularly micro-tasks (the most prevalent in the sample), are associated with the lowest levels of three ordinal income-related indicators: monthly hours dedicated to digital platform work (DPW), the proportion of total income derived from DPW, and DPW income expressed as a share of the national median.

In contrast, the right side of Dimension 1 is characterized by physically demanding, on-location activities such as delivery work (the second most prevalent task in the sample), home repair and maintenance tasks (i.e., “chores”), and transport. These activities also correspond to the higher categories of the same three income-related indicators.

The second dimension, depicted on the Y-axis, differentiates digital platform workers (DPWs) operating under self-employment arrangements from those engaged in other types of work arrangements. The latter encompass both informal arrangements (e.g., ‘No contract’), which are more common among microtask workers on the left, and formal contracts (fixed-term, permanent, or student), which are more prevalent among individuals performing high-commitment, on-location tasks on the right. Because self-employment occurs with similar frequency across both web-based and on-location tasks, it does not strongly align with this dimension.

Finally, the spread of countries across both dimensions highlights substantial cross-national variation in the composition of task types and employment conditions. These patterns may reflect underlying structural differences in labour markets, as well as recruitment strategies specific to the study. For example, the United Kingdom, Belgium, and Spain are primarily associated with remote microtasking, while approximately 70% of *delivery* workers originate from Denmark and Poland. Finland, in contrast, is more prominently represented among participants engaged in online tasks such as *writing*, *marketing*, and *administration*.

### Strengths and limitations

Several limitations must be considered when interpreting findings from the GIG-OSH cohort. First, each country implemented different recruitment strategies tailored to their context, leading to sample compositions that may disproportionately represent certain types of platform workers (e.g., mostly food couriers in one country and mostly online freelancers in another). However, as it is known, platform workers are hard to recruit because no list of platforms exists (2,22) and this population also contains a large share of undocumented migrants (23). Therefore, it is common for different recruitment strategies to be used in this type of population (16). Second, due to the different recruitment strategies and that we don’t have a population of platform workers from where we can draw a random sample, the sample is not statistically representative of the entire platform workforce in each country. However, it does not preclude the ability to detect and estimate exposure–outcome associations and extract valid inferences, assuming adequate control of confounding variables (24). No weighting or benchmarking against the labour force survey (LFS) data was possible at this stage due to limited national statistics on platform work in most countries. Third, the study faced substantial attrition between waves, with a high dropout rate and only one country (Spain) completing a third follow-up wave, which limits the longitudinal power and generalizability of long-term results. It is well known that conducting research with vulnerable or hard-to-reach populations is challenging, as they tend to have high rates of attrition and survey nonresponse, often due to a lack of trust (25,26). Fourth, all data were self-reported, which may introduce recall bias or social desirability bias, although the questionnaire was informed by validated instruments when available (27,28). Fifth, cross-national harmonization of certain variables was limited due to differences in national classification systems. Whenever possible, we used existing items from established European cross-national surveys, such as the EWCS, to enhance comparability across countries. For the remaining questions, discussions and adaptations were undertaken among the participating teams to ensure conceptual clarity and cultural relevance while maintaining maximum consistency and comparability. Finally, the study was unable to accurately capture hourly wages or waiting time between tasks, which could be critical variables for understanding economic insecurity among platform workers.

Despite these limitations, the GIG-OSH cohort presents several key strengths. Most importantly, the GIG-OSH cohort constitutes the first large-scale longitudinal effort to investigate occupational safety and health among digital platform workers across multiple European countries. Despite the inherent challenges of reaching this highly mobile and fragmented workforce, the study successfully included both web-based and on-location platform workers, allowing for comparative analyses across diverse task types, contractual arrangements, and national contexts. Most published studies focus on a single profile of these workers, predominantly on-location workers such as riders (11,29), and are usually limited to one country (30,31), or at most, two or three different countries (32). Furthermore, the cohort spans seven European countries with varying welfare state regimes and labour market regulations (13,33), offering valuable cross-national insights into the social determinants of health among platform workers. Finally, the GIG-OSH project has a mixed-methods approach (34,35) and stakeholder-informed questionnaire design that further enhances the study’s relevance and depth, capturing a wide range of occupational hazards often overlooked in traditional labour surveys, including psychosocial, algorithmic, and task-specific risks.

### Conclusions

The GIG-OSH cohort represents the first large-scale, longitudinal study examining occupational safety and health among digital platform workers across multiple European countries. By including both web-based and on-location workers, the study captures the heterogeneity of platform work and enables cross-national and cross-task comparisons of employment conditions, exposures, and health outcomes. Despite methodological challenges—including non-probability sampling, and attrition, —the study provides robust empirical insights into the working conditions and health status of a hard-to-reach and under-researched workforce. The findings highlight key differences between platform work modalities, particularly regarding employment formality, income dependency, working time, and exposure to physical and psychosocial risks. Furthermore, the GIG-OSH project adopts a holistic approach to the study of platform work by integrating quantitative and qualitative methods. Beyond combining methodological perspectives, the GIG-OSH project explicitly incorporates the viewpoints of platform workers as well as those of key stakeholders and institutional actors. This multi-actor, mixed-methods approach aims to capture the complexity of platform work and its implications for occupational safety and health.

Given the increasing policy relevance of working time and income precarity in the gig economy (36,37), future waves of the GIG-OSH study should prioritize the development of more precise tools to measure hourly earnings and unpaid waiting time. These indicators are essential for capturing the real value of labour in a task-based remuneration model, where traditional monthly income metrics may be misleading or non-comparable across work types. Furthermore, future research should aim to develop strategies to include other subgroups of platform workers that are currently underrepresented in GIG-OSH, such as web-based platform workers providing medical services, and domestic and care platform workers.

## Collaboration

The GIG-OSH consortium invites researchers to contact the corresponding author for requests for statistical code used in the multidimensional description. Requests for data availability can be submitted to the coordinators of the project TB ( and NMS and will be reviewed by the GIG-OSH steering committee.

## Participant consent

Obtained.

## Ethics approval

The cohort was approved by the local research ethics committees, and participants signed an informed consent form prior to enrolment.

Spain: Ethical approval was obtained from the Drug Research Ethics Committee (CEIm) of Hospital del Mar (reference number 2023/11095/I-).

Sweden: Ethical permission was granted for the study by the Regional Ethics Board of Stockholm with no. 2023-02230-01. Belgium, Denmark, Finland and Poland also fell under this ethical approval of Sweden.

The UK: Ethics approval was obtained from the Humanities and Social Sciences Research Ethics Committee (HSSREC), the University of Warwick (UoW) (ref.no. HSSREC 201/23-24). After the project started, the Danish data were managed from UoW and therefore also fall under UK ethics approval.

## Supporting information

Summplemental Material B

## Data Availability

All data produced in the present study are available upon reasonable request to the authors

