## Supplementary material for "Cohort profile: Description of the GIG-OSH longitudinal cohort on occupational safety and health of digital platforms workers in Europe": Summplemental Material B

### Supplemental Material B. Further Methodological Specification and Results of the Nonlinear Canonical Correlation Analysis

This supplemental material provides further methodological specification and output of the nonlinear canonical correlation analysis (NLCCA) conducted using the *homals* package in R.

Two sets of variables were used in the NLCCA analysis. Set 1 consisted of a 15-category variable capturing the main type of tasks performed by digital platform workers. These categories included on-location services (such as delivery, transport, household services, personal care, and teaching) and remote activities (including micro-tasks, administrative work, writing, creative production, marketing, professional services, and IT-related tasks), as well as residual "other" categories for both modalities.

**Table SB1.** Values of the variable 'Main task in Digital Platform Work' included in Set 1

| Value Label in Figure 2 | Question: "On which task do you spend the most amount of time" |
|---|---|
| **Delivery** | Delivery services on location (food, meals, groceries) |
| **Transport** | Transport services on location (taxi, driving, moving services) |
| **Chores** | Small chores on location (painting, electrician, plumbing, repairs, gardening) |
| **Household** | Household help on location (cleaning, cooking, babysitting, pet sitting) |
| **Personal Care** | Personal care on location (hairdressing, beauty services, home help) |
| **Teaching** | Teaching and classes on location (tutoring, lessons) |
| **Remote Micro-Tasks** | Remote micro-tasks (tagging, classification, surveys, content review) |
| **Administration** | Remote administration & data entry (customer service, transcription) |
| **Writing** | Remote writing & translation (copywriting, proofreading, translating) |
| **Creative** | Remote creative & multimedia (design, animation, photo editing) |
| **Marketing** | Remote sales & marketing support (social media, SEO, advertising) |
| **Professional** | Remote professional services (legal, bookkeeping, project management) |
| **IT** | Remote software & technology work (data science, apps, game development) |
| **Other/Unk On Location** | Other services performed on location |
| **Other/Unk Remote** | Other services performed remotely |

Set 2 included key indicators of employment conditions in digital platform work, namely type of work arrangement, monthly hours devoted to platform work, the proportion of total income derived from platform work, platform income expressed as a share of the national median income, and the existence of another job. Within this set, the variables specified as ordinal were originally measured as continuous numerical variables and were subsequently recoded into quintiles for inclusion in the nonlinear canonical correlation analysis. This recoding enabled ordinal categorisation and centroid representation in the optimal scaling solution.

**Table SB2.** Variables in the Set 2 Employment Conditions

| Variable | Description | Categories in Figure 2 | Measurement Level |
|---|---|---|---|
| **Type of work arrangement** | Employment contract or work regime | Self-Employed; Permanent; Fixed-Term; No Contract; Student Contract; Other/DK contract | Nominal |
| **Hours/month committed to DPW** | Monthly hours devoted to DPW | 0–5; 5–15; 15–30; 30–80; 80+ | Ordinal |
| **Income from DPW as % median income** | Platform income as % of national median | 2–3%; 3–5%; 5–17%; 17–39%; 39–107% | Ordinal |
| **Share of personal income from DPW (%)** | Share of total personal income from DPW | <10%; 10–25%; 26–50%; 51–75%; 76–100% | Ordinal |
| **Another job?** | Presence of another job | Another job; No other job | Binary |

In addition to the two main sets of variables included in the estimation procedure, two variables were incorporated as supplementary (passive) variables: Country and Task type/Location. These variables were specified as nominal and were not used in the estimation of object scores or category quantifications. Instead, following convergence of the alternating least squares algorithm, their category points were projected onto the solution space. This procedure allows their positions to be interpreted relative to the extracted dimensions without influencing the optimal scaling solution.

**Table SB3.** Supplementary variables

| Variable | Description | Categories in Figure 2 | Measurement Level |
|---|---|---|---|
| **Country** | Respondent country | FIN (Finland); DEN (Denmark); SWE (Sweden); BEL (Belgium); ESP (Spain); POL (Poland); UK (United Kingdom) | Nominal |
| **Task type** | Remote vs on-location work | Web-based; On location | Binary |

### Model Specification

To examine the relational structure between the two sets of variables, a NLCCA was conducted within the Gifi system of descriptive multivariate analysis (Gifi, A., 1990), utilizing the homals package in R. The model's dimensionality was determined through an iterative process evaluating solutions of increasing dimensionality. A three-dimensional solution was ultimately selected as the optimal trade-off between mathematical stability and theoretical parsimony as indicated by convergence stability and interpretability of the resulting dimensions. This specification resolves the iterative instabilities (specifically the non-monotonic loss function behavior) observed in lower-dimensional solutions, providing sufficient geometric space to accommodate the high cardinality of the task-related variables.

### Global Fit and Interpretation of Loss

The finalized model achieved a remarkably low loss value ($1.31\times10-5$) after 3 iterations. While such a value might typically suggest an over-fitted model, in the context of the homals algorithm and the inclusion of a variable with 15 categories, it is interpreted here as an indicator of perfect convergence rather than a perfect correlation: this low loss confirms that the Alternating Least Squares (ALS) algorithm successfully found a stable state where the distances between the quantified objects and their respective category centroids were minimized.

### Eigenvalues and Dimensional Significance

In contrast to the convergence indicated by the loss function, the eigenvalues ($\lambda$) provide a more substantive measure of the discriminatory power and internal consistency of each axis. The first and second dimensions yielded modest eigenvalues of 0.157 and 0.191, respectively. Interpretation clues for the two first axes have already been provided in the main text. The emergence of a significant third dimension ($\lambda$=0.154) further underscores the complexity of the digital platform labor ecosystem. This third axis accounts for a distinct, orthogonal portion of the relational structure—specifically identifying strategies of pluriactivity—that operates independently of the legal-contractual on location / web based echelons defined by the first two axes.

Please note that, the eigenvalues in a HOMALS/Gifi framework must not be interpreted as the percentage of total variance explained in the sense of a classical Principal Component Analysis (PCA). In PCA, eigenvalues represent the decomposition of a fixed variance-covariance matrix in descending order of importance. In contrast, HOMALS eigenvalues represent the mean of the discrimination measures for all variables within a specific dimension.

*It is important to note that, for the purposes of visual representation in Figure 2, these dimensions have been transposed: Dimension 2 (named 1 in the graph) —which exhibits the highest degree of homogeneity and structural consistency—is represented on the abscissa (x-axis), while Dimension 1 (named 2 in the graph) is represented on the ordinate (y-axis)*.

### Variable loadings

Table SB4 presents the variable loadings obtained from the nonlinear canonical correlation analysis. These loadings indicate the degree of association between each optimally scaled variable and the extracted dimensions. The magnitude and pattern of the loadings justify the substantive interpretation assigned to the axes, as variables with higher loadings contribute more strongly to the definition of each dimension.

**Table SB4. Variable loadings**

| Variable | Dim 1 | Dim 2 | Dim 3 |
|---|---|---|---|
| Main Task (15 categories) | -0,212 | -0,552 | -0,258 |
| Contract Type | **0,546** | 0,03 | 0,116 |
| Secondary Job | 0,115 | 0,237 | **-0,297** |
| Earnings (quintiles) | 0,138 | **0,547** | 0,128 |
| Work Hours (quintiles) | 0,083 | **0,486** | 0,096 |
| Platform Income Prop. (quintiles) | 0,198 | **0,419** | -0,093 |
| *Country* | 0,111 | -0,339 | -0,141 |
| *Main Task (Binary)* | 0,072 | **-0,577** | -0,121 |

[1]Bold values indicate the primary contributing variables for each dimension.

### Category quantifications

Table SB5 reports category centroids obtained from the nonlinear canonical correlation analysis. Centroids represent the coordinates of category points in the optimally scaled solution space and were used for graphical representation in Figure 2.

**Table SB5.** Category Quantifications (Centroids) for the 3-D HOMALS Solution

| Variable | Category | Dimension | | |
|---|---|---|---|---|
| | | D1 | D2 | D3 |
| **Main Task** | Administration | 0,00534283 | -0,00823828 | 0,01251417 |
| | Chores | -0,01134069 | 0,00816343 | 0,02121698 |

| | | | | |
|---|---|---|---|---|
| | Creative | 0,00724614 | -0,00554590 | 0,00983271 |
| | Delivery | 0,00246596 | 0,01461938 | -0,00354028 |
| | Household | -0,01168069 | 0,00099707 | 0,01540162 |
| | IT | 0,00790973 | -0,00326726 | 0,00846122 |
| | Marketing | 0,00857210 | -0,00735125 | 0,01001856 |
| | Other/Unk non loc. | -0,00185028 | 0,01206767 | 0,01307015 |
| | Other/Unk Remote | 0,00077995 | -0,01199460 | -0,00075259 |
| | Personal Care | -0,01136404 | -0,00177301 | 0,02210526 |
| | Professional | 0,00720123 | 0,00048252 | 0,01836011 |
| | Remote Micro-tasks | -0,00827824 | -0,01101278 | -0,00908722 |
| | Teaching | -0,00391978 | 0,00154966 | 0,00686395 |
| | Transport | -0,01354515 | 0,00967719 | 0,01921712 |
| | Writing | 0,02336023 | -0,00798562 | 0,00213858 |
| --- | --- | --- | --- | --- |
| **Contract Type** | Fixed-Term | -0,00435935 | 0,00716856 | 0,01452249 |
| | No Contract | -0,00743610 | -0,00476876 | -0,00422641 |
| | Other/DK Contract | 0,00197915 | -0,00335893 | -0,00475518 |
| | Permanent | -0,00788292 | 0,01589549 | 0,00423307 |
| | Self-Employed | 0,01515089 | -0,00038036 | 0,00212406 |
| | Student contract | -0,00215359 | 0,01090827 | 0,00456661 |
| --- | --- | --- | --- | --- |
| **Another Job** | Another job | -0,00157970 | -0,00325366 | 0,00407622 |
| | No other job | 0,00267962 | 0,00551916 | -0,00691445 |
| --- | --- | --- | --- | --- |
| **Earnings Deciles** | (a) 2-3% | -0,00440325 | -0,01050571 | -0,00540498 |
| | (b) 3-5% | 0,00006979 | -0,01154491 | -0,00463734 |
| | (c) 5-17% | -0,00064432 | 0,00266751 | 0,00877578 |
| | (d) 17-39% | 0,00183931 | 0,00875851 | 0,00648367 |
| | (e) 39-107% | 0,00371779 | 0,01357512 | -0,00468703 |
| --- | --- | --- | --- | --- |
| **Workhours Dec.** | (a) 0-5 hrs. | -0,00345829 | -0,01000453 | -0,00863219 |
| | (b) 5-15 hrs. | -0,00071894 | -0,00661940 | 0,00045778 |
| | (c) 15-30 hrs. | 0,00234630 | -0,00364506 | 0,00807981 |
| | (d) 30-80 hrs. | 0,00060065 | 0,00607485 | 0,00484329 |
| | (e) 80+ hrs. | 0,00129637 | 0,01413678 | -0,00452404 |
| --- | --- | --- | --- | --- |
| **Inc. Platform** | < 10% | -0,00277436 | -0,00841820 | -0,00291991 |
| | 10-25% | -0,00150318 | 0,00062999 | 0,00858224 |
| | 26-50% | 0,00049471 | 0,00658273 | 0,00680012 |
| | 51-75% | -0,00075135 | 0,00692059 | 0,01036287 |

| | | | | |
|---|---|---|---|---|
| | 76-100% | 0,00740389 | 0,01056930 | -0,01030583 |
| --- | --- | --- | --- | --- |
| **Country** | BEL | -0,00282355 | -0,00454926 | -0,00263725 |
| | DEN | 0,00326203 | 0,00791947 | -0,00375500 |
| | ESP | -0,00187074 | -0,00315561 | -0,00351837 |
| | FIN | 0,01183467 | -0,00689272 | 0,00119885 |
| | POL | -0,00384492 | 0,00806324 | 0,00727853 |
| | SWE | 0,00061849 | -0,00303754 | -0,00068634 |
| | UK | -0,00432284 | -0,00694051 | -0,00154967 |
| --- | --- | --- | --- | --- |
| **Task Binary** | On location | -0,00145104 | 0,01165425 | 0,00244720 |
| | Web-based | 0,00113715 | -0,00913315 | -0,00191781 |

### R code

```
names(variables_for_analysis)

[1]
"main_task2"                    "Contract_type2"               "ano
ther_job2"

[4]
"earnings_quintiles"            "workhours_quintiles"

[6]
"prop_income_quintiles"  "countryshort"  "main_task_binary"



sets <-list(c(1), c(2,3,4,5,6),c(7,8))  # Definition of sets

level_var<-
c("nominal","nominal","nominal","ordinal","ordinal","ordinal","n
ominal","nominal") # measurement level of the variables

homals_result <- homals::homals(variables_for_analysis,

                        ndim =3,

                        sets = sets, # Define the sets of
variables

                        verbose=T,

           level = level_var,
```

```
                        active =
c(rep(TRUE,6),c(FALSE,FALSE)))  # "countryshort"  &
"main_task_binary"are defined as supplementary variables
```
