## Supplemental material A for "Cohort profile: Description of the GIG-OSH longitudinal cohort on occupational safety and health of digital platforms workers in Europe"

**Supplementary material A. Supplementary tables.**

**Table SA1.** Variables included in the different constructed composite indices.

| Index | Definition | Variables included | Scale and interpretation |
|---|---|---|---|
| **Earnings index** | Reflects normalised hourly wages relative to the country-specific median | Country income variables, monthly working hours | 0-100; higher values = higher relative earnings |
| **Contract type index** | Captures the degree of employment formality | Contract type | 0-100; higher values = more formal arrangements |
| **Working time quality index** | Assesses the quality of working time (predictability, duration, rest) | Unsocial working hours, work-life balance, unpaid working hours, monthly working hours | 0-100; higher values = better working time quality |
| **Work intensity index** | Evaluates workload intensity and pressure | Risk of dealing with difficult clients, risk of emotionally disturbing situations, working at very high speed | 0-100; higher values = greater intensity |
| **Physical environment index** | Reflects exposure to physical risks in the workplace | Exposure to vibration, exposure to noise, exposure to high or low temperatures, exposure to vapours or chemical substances, repetitive hand or arm movements, tiring or painful positions, carrying or moving heavy loads, working at heights, exposure to traffic risks, exposure to adverse weather, prolonged sitting, risk of fraud or theft | 0-100; higher values = better physical environment |
| **Social environment index** | Assesses social relations and support at work | Working alone, experience of discrimination, experience of harassment, risk of physical attack | 0-100; higher values = better social environment |
| **Autonomy index** | Measures workers' control over their tasks and methods | Control over the order of tasks, control over methods of work, control over speed or rate of work | 0-100; higher values = greater autonomy |

**Table SA2.** Baseline (n=3,945) and follow-up (n=385) sociodemographic and labour characteristics of the 3,945 participants in the GIG-OSH cohort by country.

| | Baseline | | | | | | | Followup | | | | | | |
|---|---|---|---|---|---|---|---|---|---|---|---|---|---|---|
| | Belgium | Denmark | Finland | Poland | Spain | Sweden | UK | Belgium | Denmark | Finland | Poland | Spain | Sweden | UK |
| | N = 280 | N = 718 | N = 456 | N = 858 | N = 575 | N = 546 | N = 512 | N = 16 | N = 43 | N = 54 | N = 55 | N = 137 | N = 54 | N = 26 |
| | n (%) | n (%) | n (%) | n (%) | n (%) | n (%) | n (%) | n (%) | n (%) | n (%) | n (%) | n (%) | n (%) | n (%) |
| **Sociodemographic variables** | | | | | | | | | | | | | | |
| **Gender** | | | | | | | | | | | | | | |
| Male | 147 (53.07) | 557 (77.68) | 251 (56.92) | 439 (51.29) | 310 (54.29) | 383 (70.79) | 215 (42.24) | 11 (73.33) | 36 (83.72) | 26 (48.15) | 29 (52.73) | 64 (47.41) | 37 (68.52) | 14 (56.00) |
| Female | 125 (45.13) | 148 (20.64) | 181 (41.04) | 403 (47.08) | 252 (44.13) | 154 (28.47) | 286 (56.19) | 4 (26.67) | 7 (16.28) | 27 (50.00) | 26 (47.27) | 68 (50.37) | 16 (29.63) | 11 (44.00) |
| Other | 4 (1.44) | 9 (1.26) | 7 (1.59) | 6 (0.70) | 9 (1.58) | 3 (0.55) | 6 (1.18) | 0 (0.00) | 0 (0.00) | 1 (1.85) | 0 (0.00) | 3 (2.22) | 1 (1.85) | 0 (0.00) |
| I prefer not to say | 1 (0.36) | 3 (0.42) | 2 (0.45) | 8 (0.93) | 0 (0.00) | 1 (0.18) | 2 (0.39) | 0 (0.00) | 0 (0.00) | 0 (0.00) | 0 (0.00) | 0 (0.00) | 0 (0.00) | 0 (0.00) |
| Missing | 3 | 1 | 15 | 2 | 4 | 5 | 3 | 1 | 0 | 0 | 0 | 2 | 0 | 1 |
| **Age (mean, SD)** | 34.41 (13.61) | 32.38 (9.80) | 32.25 (9.16) | 30.68 (10.14) | 33.75 (10.94) | 31.28 (8.48) | 35.77 (11.02) | 35.93 (12.85) | 34.15 (12.01) | 35.73 (9.01) | 36.62 (10.54) | 37.67 (11.58) | 33.47 (9.02) | 39.00 (12.82) |
| Missing | 10 | 51 | 27 | 3 | 10 | 17 | 15 | 1 | 4 | 2 | 0 | 2 | 1 | 0 |
| **Migration** | | | | | | | | | | | | | | |
| No-immigrant | 177 (63.9) | 257 (36.25) | 281 (63.57) | 813 (97.25) | 342 (60.64) | 243 (45.25) | 344 (69.08) | 13 (81.25) | 21 (48.84) | 35 (64.81) | 52 (100.00) | 87 (64.93) | 29 (54.72) | 20 (83.33) |
| Immigrant | 95 (34.3) | 428 (60.37) | 150 (33.94) | 20 (2.39) | 213 (37.77) | 277 (51.58) | 132 (26.51) | 3 (18.75) | 22 (51.16) | 19 (35.19) | 0 (0.00) | 46 (34.33) | 24 (45.28) | 4 (16.67) |
| I prefer not to say | 5 (1.81) | 24 (3.39) | 11 (2.49) | 3 (0.36) | 9 (1.60) | 17 (3.17) | 22 (4.42) | 0 (0.00) | 0 (0.00) | 0 (0.00) | 0 (0.00) | 1 (0.75) | 0 (0.00) | 0 (0.00) |
| Missing | 5 | 9 | 14 | 22 | 11 | 9 | 14 | 0 | 0 | 0 | 3 | 3 | 1 | 2 |
| **Education** | | | | | | | | | | | | | | |
| Primary | 7 (2.53) | 1 (0.14) | 30 (6.77) | 27 (3.17) | 12 (2.09) | 17 (3.17) | 6 (1.18) | 0 (0.00) | 0 (0.00) | 1 (1.85) | 0 (0.00) | 2 (1.47) | 3 (5.56) | 0 (0.00) |
| Secondary | 66 (23.83) | 277 (39.24) | 132 (29.80) | 525 (61.62) | 169 (29.49) | 134 (24.95) | 152 (29.80) | 5 (31.25) | 19 (44.19) | 10 (18.52) | 27 (49.09) | 37 (27.21) | 17 (31.48) | 8 (30.77) |

| | | | | | | | | | | | | | | |
|---|---|---|---|---|---|---|---|---|---|---|---|---|---|---|
| Tertiary | 192 (69.31) | 393 (55.67) | 276 (62.30) | 299 (35.09) | 392 (68.41) | 263 (48.98) | 343 (67.25) | 10 (62.50) | 23 (53.49) | 42 (77.78) | 28 (50.91) | 97 (71.32) | 19 (35.19) | 18 (69.23) |
| Unrecognized | 12 (4.33) | 35 (4.96) | 5 (1.13) | 1 (0.12) | 0 (0.00) | 123 (22.91) | 9 (1.76) | 1 (6.25) | 1 (2.33) | 1 (1.85) | 0 (0.00) | 0 (0.00) | 15 (27.78) | 0 (0.00) |
| Missing | 3 | 12 | 13 | 6 | 2 | 9 | 2 | 0 | 0 | 0 | 0 | 1 | 0 | 0 |
| **Ends meet** | | | | | | | | | | | | | | |
| Very easily | 31 (11.19) | 57 (8.20) | 31 (7.01) | 74 (8.76) | 46 (8.07) | 41 (7.65) | 32 (6.46) | 4 (25.00) | 4 (9.30) | 4 (7.41) | 0 (0.00) | 17 (12.59) | 4 (7.41) | 3 (12.50) |
| Easily | 64 (23.10) | 115 (16.55) | 77 (17.42) | 116 (13.73) | 103 (18.07) | 85 (15.86) | 93 (18.79) | 3 (18.75) | 11 (25.58) | 8 (14.81) | 9 (16.67) | 20 (14.81) | 10 (18.52) | 7 (29.17) |
| Fairly easily | 78 (28.16) | 151 (21.73) | 123 (27.83) | 271 (32.07) | 128 (22.46) | 162 (30.22) | 188 (37.98) | 3 (18.75) | 8 (18.60) | 12 (22.22) | 17 (31.48) | 39 (28.89) | 18 (33.33) | 6 (25.00) |
| With some difficulty | 60 (21.66) | 214 (30.79) | 117 (26.47) | 254 (30.06) | 190 (33.33) | 147 (27.43) | 115 (23.23) | 3 (18.75) | 11 (25.58) | 19 (35.19) | 19 (35.19) | 40 (29.63) | 14 (25.93) | 5 (20.83) |
| With difficulty | 26 (9.39) | 78 (11.22) | 56 (12.67) | 60 (7.10) | 56 (9.82) | 52 (9.70) | 33 (6.67) | 1 (6.25) | 3 (6.98) | 5 (9.26) | 5 (9.26) | 9 (6.67) | 2 (3.70) | 2 (8.33) |
| With great difficulty | 9 (3.25) | 51 (7.34) | 28 (6.33) | 33 (3.91) | 39 (6.84) | 38 (7.09) | 24 (4.85) | 2 (12.50) | 6 (13.95) | 4 (7.41) | 2 (3.70) | 9 (6.67) | 5 (9.26) | 1 (4.17) |
| I prefer not to say | 9 (3.25) | 29 (4.17) | 10 (2.26) | 37 (4.38) | 8 (1.40) | 11 (2.05) | 10 (2.02) | 0 (0.00) | 0 (0.00) | 2 (3.70) | 2 (3.70) | 1 (0.74) | 1 (1.85) | 0 (0.00) |
| Missing | 3 | 23 | 14 | 13 | 5 | 10 | 17 | 0 | 0 | 0 | 1 | 2 | 0 | 2 |
| **Age** | 34.41 (13.61) | 32.38 (9.80) | 32.25 (9.16) | 30.68 (10.14) | 33.75 (10.94) | 31.28 (8.48) | 35.77 (11.02) | 35.93 (12.85) | 34.15 (12.01) | 35.73 (9.01) | 36.62 (10.54) | 37.67 (11.58) | 33.47 (9.02) | 39.00 (12.82) |
| Missing | 10 | 51 | 27 | 3 | 10 | 17 | 15 | 1 | 4 | 2 | 0 | 2 | 1 | 0 |
| **Labour characteristics** | | | | | | | | | | | | | | |
| **Platform task** | | | | | | | | | | | | | | |
| On location | 112 (40.00) | 576 (80.22) | 124 (27.19) | 631 (73.54) | 186 (32.35) | 197 (36.08) | 122 (23.83) | 7 (58.33) | 26 (89.66) | 6 (15.38) | 17 (54.84) | 20 (16.13) | 11 (26.19) | 10 (41.67) |
| Web-based | 168 (60.00) | 141 (19.64) | 332 (72.81) | 227 (26.46) | 388 (67.48) | 349 (63.92) | 390 (76.17) | 5 (41.67) | 3 (10.34) | 33 (84.62) | 14 (45.16) | 104 (83.87) | 31 (73.81) | 14 (58.33) |
| Unknown | 0 (0.00) | 1 (0.14) | 0 (0.00) | 0 (0.00) | 1 (0.17) | 0 (0.00) | 0 (0.00) | 0 (0.00) | 0 (0.00) | 0 (0.00) | 0 (0.00) | 0 (0.00) | 0 (0.00) | 0 (0.00) |
| Missing | 0 | 0 | 0 | 0 | 0 | 0 | 0 | 4 | 14 | 15 | 24 | 13 | 12 | 2 |
| **Contract type** | | | | | | | | | | | | | | |
| Without | 146 (56.37) | 148 (25.26) | 107 (27.30) | 259 (33.59) | 324 (58.70) | 214 (45.34) | 290 (60.29) | 6 (50.00) | 6 (21.43) | 13 (34.21) | 9 (29.03) | 86 (69.35) | 31 (73.81) | 11 (45.83) |

| | | | | | | | | | | | | | | |
|---|---|---|---|---|---|---|---|---|---|---|---|---|---|---|
| Permanent | 7 (2.70) | 77 (13.14) | 12 (3.06) | 193 (25.03) | 31 (5.62) | 32 (6.78) | 17 (3.53) | 0 (0.00) | 4 (14.29) | 1 (2.63) | 5 (16.13) | 8 (6.45) | 2 (4.76) | 2 (8.33) |
| Temporal | 5 (1.93) | 17 (2.90) | 23 (5.87) | 119 (15.43) | 23 (4.17) | 44 (9.32) | 21 (4.37) | 2 (16.67) | 2 (7.14) | 2 (5.26) | 4 (12.90) | 3 (2.42) | 2 (4.76) | 4 (16.67) |
| Self-employed | 54 (20.85) | 236 (40.27) | 219 (55.87) | 116 (15.05) | 137 (24.82) | 115 (24.36) | 109 (22.66) | 2 (16.67) | 12 (42.86) | 21 (55.26) | 11 (35.48) | 26 (20.97) | 5 (11.90) | 6 (25.00) |
| Student contract | 11 (4.25) | 25 (4.27) | 5 (1.28) | 49 (6.36) | 0 (0.00) | 11 (2.33) | 3 (0.62) | 1 (8.33) | 0 (0.00) | 0 (0.00) | 0 (0.00) | 0 (0.00) | 1 (2.38) | 0 (0.00) |
| Other | 2 (0.77) | 2 (0.34) | 0 (0.00) | 1 (0.13) | 1 (0.18) | 2 (0.42) | 0 (0.00) | 1 (8.33) | 1 (3.57) | 0 (0.00) | 2 (6.45) | 0 (0.00) | 0 (0.00) | 0 (0.00) |
| I don't know | 34 (13.13) | 81 (13.82) | 26 (6.63) | 34 (4.41) | 36 (6.52) | 54 (11.44) | 41 (8.52) | 0 (0.00) | 3 (10.71) | 1 (2.63) | 0 (0.00) | 1 (0.81) | 1 (2.38) | 1 (4.17) |
| Missing | 21 | 132 | 64 | 87 | 23 | 74 | 31 | 4 | 15 | 16 | 24 | 13 | 12 | 2 |
| **Another job** | | | | | | | | | | | | | | |
| Yes | 174 (63.04) | 376 (52.59) | 240 (53.33) | 558 (65.96) | 341 (59.72) | 317 (58.49) | 359 (70.81) | - | - | - | - | - | - | - |
| No | 102 (36.96) | 339 (47.41) | 210 (46.67) | 288 (34.04) | 230 (40.28) | 225 (41.51) | 148 (29.19) | - | - | - | - | - | - | - |
| Missing | 4 | 3 | 6 | 12 | 4 | 4 | 5 | - | - | - | - | - | - | - |
| **Months as gig worker** | | | | | | | | | | | | | | |
| < 1 month | 33 (12.00) | 73 (10.58) | 58 (13.46) | 71 (8.78) | 30 (5.25) | 81 (15.43) | 33 (6.56) | - | - | - | - | - | - | - |
| 1-3 months | 33 (12.00) | 106 (15.36) | 87 (20.19) | 198 (24.47) | 62 (10.86) | 122 (23.24) | 62 (12.33) | - | - | - | - | - | - | - |
| 3-6 months | 19 (6.91) | 107 (15.51) | 50 (11.60) | 186 (22.99) | 70 (12.26) | 69 (13.14) | 40 (7.95) | - | - | - | - | - | - | - |
| 6-12 months | 18 (6.55) | 82 (11.88) | 45 (10.44) | 147 (18.17) | 52 (9.11) | 55 (10.48) | 50 (9.94) | - | - | - | - | - | - | - |
| > 12 months | 172 (62.55) | 322 (46.67) | 191 (44.32) | 207 (25.59) | 357 (62.52) | 198 (37.71) | 318 (63.22) | - | - | - | - | - | - | - |
| Missing | 5 | 28 | 25 | 49 | 4 | 21 | 9 | - | - | - | - | - | - | - |
| **Income as % of median (mean, SD)** | 13.37 (19.04) | 29.07 (26.76) | 19.52 (23.78) | 25.40 (16.86) | 18.12 (22.36) | 20.62 (22.28) | 9.71 (15.95) | 12.56 (19.60) | 39.47 (27.79) | 22.21 (25.99) | 24.08 (18.30) | 17.51 (23.59) | 10.16 (11.85) | 9.55 (10.70) |
| Missing | 28 | 106 | 53 | 107 | 27 | 58 | 46 | 5 | 16 | 19 | 24 | 15 | 14 | 5 |
| **Total hours (mean, SD)** | 34.23 (65.95) | 61.46 (72.70) | 33.63 (49.72) | 68.90 (70.43) | 60.10 (93.77) | 36.59 (58.12) | 32.35 (55.28) | 20.71 (19.90) | 78.09 (62.15) | 46.81 (59.73) | 53.87 (49.75) | 42.53 (63.73) | 29.21 (37.24) | 113.21 (151.00) |
| Missing | 36 | 167 | 82 | 108 | 31 | 91 | 47 | 4 | 15 | 17 | 24 | 13 | 12 | 2 |

**Table SA3.** Baseline (n=3,945) and follow-up (n=385) occupational safety and health (OSH) conditions, self-perceived health, and work environment characteristics of participants in the GIG-OSH cohort by country.

| | Baseline | | | | | | | Followup | | | | | | |
|---|---|---|---|---|---|---|---|---|---|---|---|---|---|---|
| | Belgium | Denmark | Finland | Poland | Spain | Sweden | UK | Belgium | Denmark | Finland | Poland | Spain | Sweden | UK |
| | N = 280 | N = 718 | N = 456 | N = 858 | N = 575 | N = 546 | N = 512 | N = 16 | N = 43 | N = 54 | N = 55 | N = 137 | N = 54 | N = 26 |
| **Occupational safety and health (OSH) conditions, self-perceived health, and work environment characteristics** | n (%) | n (%) | n (%) | n (%) | n (%) | n (%) | n (%) | n (%) | n (%) | n (%) | n (%) | n (%) | n (%) | n (%) |
| **OSH information** | | | | | | | | | | | | | | |
| Very well informed | 63 (24.14) | 140 (23.26) | 67 (16.83) | 199 (25.51) | 117 (21.20) | 113 (23.59) | 161 (33.54) | - | - | - | - | - | - | - |
| Well informed | 111 (42.53) | 243 (40.37) | 148 (37.19) | 372 (47.69) | 251 (45.47) | 196 (40.92) | 204 (42.50) | - | - | - | - | - | - | - |
| Not very well informed | 49 (18.77) | 150 (24.92) | 119 (29.90) | 145 (18.59) | 106 (19.20) | 128 (26.72) | 78 (16.25) | - | - | - | - | - | - | - |
| Not at all informed | 38 (14.56) | 69 (11.46) | 64 (16.08) | 64 (8.21) | 78 (14.13) | 42 (8.77) | 37 (7.71) | - | - | - | - | - | - | - |
| Missing | 19 | 116 | 58 | 78 | 23 | 67 | 32 | - | - | - | - | - | - | - |
| **Protective equipment** | | | | | | | | | | | | | | |
| Yes, provided and paid by platform | 11 (4.23) | 73 (12.17) | 23 (5.76) | 179 (22.89) | 30 (5.41) | 32 (6.69) | 24 (5.01) | - | - | - | - | - | - | - |
| Yes, provided by platform and paid by me | 8 (3.08) | 132 (22.00) | 12 (3.01) | 145 (18.54) | 19 (3.42) | 32 (6.69) | 14 (2.92) | - | - | - | - | - | - | - |
| Yes, provided and paid by me | 39 (15.00) | 141 (23.50) | 39 (9.77) | 173 (22.12) | 345 (62.16) | 36 (7.53) | 51 (10.65) | - | - | - | - | - | - | - |
| No, I don't use | 193 (74.23) | 243 (40.50) | 308 (77.19) | 261 (33.38) | 151 (27.21) | 367 (76.78) | 381 (79.54) | - | - | - | - | - | - | - |
| Other | 9 (3.46) | 11 (1.83) | 17 (4.26) | 24 (3.07) | 10 (1.80) | 11 (2.30) | 9 (1.88) | - | - | - | - | - | - | - |
| Missing | 20 | 118 | 57 | 76 | 20 | 68 | 33 | - | - | - | - | - | - | - |
| **General health** | | | | | | | | | | | | | | |

| | | | | | | | | | | | | | | |
|---|---|---|---|---|---|---|---|---|---|---|---|---|---|---|
| Excellent | 49 (18.70) | 144 (23.88) | 71 (17.62) | 84 (10.71) | 98 (17.66) | 82 (17.08) | 82 (16.91) | 4 (25.00) | 12 (29.27) | 9 (18.00) | 2 (3.70) | 14 (10.29) | 11 (20.37) | 3 (11.54) |
| Very good | 117 (44.66) | 230 (38.14) | 156 (38.71) | 300 (38.27) | 228 (41.08) | 179 (37.29) | 184 (37.94) | 9 (56.25) | 10 (24.39) | 18 (36.00) | 15 (27.78) | 63 (46.32) | 24 (44.44) | 9 (34.62) |
| Good | 74 (28.24) | 157 (26.04) | 123 (30.52) | 312 (39.80) | 189 (34.05) | 164 (34.17) | 156 (32.16) | 2 (12.50) | 15 (36.59) | 15 (30.00) | 23 (42.59) | 45 (33.09) | 16 (29.63) | 7 (26.92) |
| Fair | 21 (8.02) | 60 (9.95) | 50 (12.41) | 79 (10.08) | 37 (6.67) | 46 (9.58) | 53 (10.93) | 0 (0.00) | 4 (9.76) | 6 (12.00) | 10 (18.52) | 14 (10.29) | 3 (5.56) | 4 (15.38) |
| Poor | 1 (0.38) | 12 (1.99) | 3 (0.74) | 9 (1.15) | 3 (0.54) | 9 (1.88) | 10 (2.06) | 1 (6.25) | 0 (0.00) | 2 (4.00) | 4 (7.41) | 0 (0.00) | 0 (0.00) | 3 (11.54) |
| Missing | 18 | 115 | 53 | 74 | 20 | 66 | 27 | 0 | 2 | 4 | 1 | 1 | 0 | 0 |
| **Health symptoms (mean, SD)** | 1.41 (1.39) | 1.22 (1.45) | 1.31 (1.43) | 1.79 (1.59) | 1.53 (1.44) | 1.27 (1.38) | 1.34 (1.47) | 1.94 (1.57) | 1.24 (1.59) | 2.34 (1.60) | 2.65 (1.63) | 1.89 (1.49) | 1.83 (1.44) | 2.31 (1.74) |
| Missing | 0 | 0 | 0 | 0 | 0 | 0 | 0 | 0 | 2 | 4 | 1 | 1 | 1 | 0 |
| **WHO-5 (mean, SD)** | 58.35 (20.71) | 58.35 (22.57) | 62.31 (19.42) | 57.89 (20.33) | 58.15 (20.28) | 57.09 (20.84) | 59.70 (20.43) | 59.25 (21.72) | 59.10 (22.20) | 56.16 (18.93) | 53.41 (19.35) | 57.62 (20.37) | 59.40 (20.48) | 60.00 (21.85) |
| Missing | 26 | 139 | 65 | 82 | 25 | 78 | 37 | 0 | 3 | 4 | 1 | 1 | 1 | 0 |
| **AMP index: automated direction systems (mean, SD)** | 55.98 (20.10) | 60.75 (18.58) | 50.84 (18.28) | 59.88 (20.70) | 62.35 (21.00) | 56.84 (18.04) | 60.68 (19.55) | - | - | - | - | - | - | - |
| Missing | 26 | 139 | 71 | 119 | 33 | 81 | 41 | - | - | - | - | - | - | - |
| **AMP index: automated evaluation systems (mean, SD)** | 44.73 (29.70) | 34.55 (31.30) | 49.17 (24.65) | 51.12 (23.52) | 46.21 (29.27) | 50.77 (28.66) | 42.88 (29.71) | - | - | - | - | - | - | - |
| Missing | 43 | 195 | 97 | 141 | 55 | 127 | 66 | - | - | - | - | - | - | - |
| **Earnings index (mean, SD)** | 1.64 (6.80) | 1.14 (4.41) | 1.07 (2.50) | 0.97 (2.25) | 0.92 (2.43) | 1.38 (3.87) | 1.08 (2.83) | - | - | - | - | - | - | - |
| Missing | 58 | 222 | 128 | 179 | 50 | 127 | 79 | - | - | - | - | - | - | - |
| **Contract type index (mean, SD)** | 13.77 (24.27) | 31.34 (33.16) | 26.45 (23.37) | 44.57 (40.67) | 16.67 (27.32) | 22.67 (30.90) | 14.41 (24.91) | - | - | - | - | - | - | - |
| Missing | 21 | 132 | 64 | 87 | 23 | 74 | 31 | - | - | - | - | - | - | - |
| **Working time quality index (mean, SD)** | 67.77 (12.41) | 63.22 (14.40) | 65.93 (11.71) | 62.62 (10.64) | 63.29 (15.47) | 64.01 (11.86) | 67.97 (11.89) | - | - | - | - | - | - | - |
| Missing | 27 | 143 | 68 | 101 | 27 | 78 | 34 | - | - | - | - | - | - | - |
| **Work intensity index (mean, SD)** | 28.48 (19.99) | 37.73 (23.78) | 32.50 (18.60) | 41.63 (20.86) | 34.20 (20.45) | 31.19 (19.25) | 30.50 (20.11) | - | - | - | - | - | - | - |

| | | | | | | | | | | | | | | |
|---|---|---|---|---|---|---|---|---|---|---|---|---|---|---|
| Missing | 15 | 102 | 51 | 53 | 15 | 58 | 27 | - | - | - | - | - | - | - |
| **Physical environment index (mean, SD)** | 20.80 (18.98) | 33.58 (19.06) | 16.01 (15.75) | 32.02 (18.29) | 16.81 (17.25) | 19.77 (17.36) | 17.12 (15.92) | - | - | - | - | - | - | - |
| Missing | 11 | 89 | 45 | 46 | 14 | 48 | 24 | - | - | - | - | - | - | - |
| **Social environment index (mean, SD)** | 76.65 (8.61) | 73.37 (11.51) | 74.19 (10.03) | 73.66 (12.55) | 74.92 (9.92) | 74.97 (10.31) | 75.92 (9.76) | - | - | - | - | - | - | - |
| Missing | 18 | 121 | 58 | 89 | 23 | 68 | 35 | - | - | - | - | - | - | - |
| **Autonomy index (mean, SD)** | 59.78 (29.26) | 56.79 (30.60) | 63.62 (23.85) | 52.37 (23.80) | 51.05 (28.02) | 60.18 (24.19) | 59.14 (27.83) | - | - | - | - | - | - | - |
| Missing | 67 | 233 | 133 | 157 | 82 | 142 | 96 | - | - | - | - | - | - | - |
